# The Neurodegenerative Disease Knowledge Portal: Propelling Discovery Through the Sharing of Neurodegenerative Disease Genomic Resources

**DOI:** 10.1101/2024.05.27.24307990

**Authors:** Allison A. Dilliott, Maria C. Costanzo, Sara Bandres-Ciga, Cornelis Blauwendraat, Bradford Casey, Quy Hoang, Hirotaka Iwaki, Dongkeun Jang, Jonggeol Jeffrey Kim, Hampton L. Leonard, Kristin S. Levine, Mary Makarious, Trang T. Nguyen, Guy A. Rouleau, Andrew B. Singleton, Patrick Smadbeck, J Solle, Dan Vitale, Mike A. Nalls, Jason Flannick, Noël P. Burtt, Sali M.K. Farhan

## Abstract

Although large-scale genetic association studies have proven useful for the delineation of neurodegenerative disease processes, we still lack a full understanding of the pathological mechanisms of these diseases, resulting in few appropriate treatment options and diagnostic challenges. To mitigate these gaps, the Neurodegenerative Disease Knowledge Portal (NDKP) was created as an open-science initiative with the aim to aggregate, enable analysis, and display all available genomic datasets of neurodegenerative disease, while protecting the integrity and confidentiality of the underlying datasets. The portal contains 218 genomic datasets, including genotyping and sequencing studies, of individuals across ten different phenotypic groups, including neurological conditions such as Alzheimer’s disease, amyotrophic lateral sclerosis, Lewy body dementia, and Parkinson’s disease. In addition to securely hosting large genomic datasets, the NDKP provides accessible workflows and tools to effectively utilize the datasets and assist in the facilitation of customized genomic analyses. Here, we summarize the genomic datasets currently included within the portal, the bioinformatics processing of the datasets, and the variety of phenotypes captured. We also present example use-cases of the various user interfaces and integrated analytic tools to demonstrate their extensive utility in enabling the extraction of high-quality results at the source, for both genomics experts and those in other disciplines. Overall, the NDKP promotes open-science and collaboration, maximizing the potential for discovery from the large-scale datasets researchers and consortia are expending immense resources to produce and resulting in reproducible conclusions to improve diagnostic and therapeutic care for neurodegenerative disease patients.

## Introduction

Neurodegenerative diseases are clinically heterogeneous and complex disorders. Given their relatively high estimates of heritability^1–4^, large-scale association studies are particularly useful for gaining a greater understanding of the pathologic mechanisms driving neurodegenerative disease processes, as demonstrated by their discoveries across neurodegenerative conditions, including in Alzheimer’s disease, amyotrophic lateral sclerosis (ALS), and Parkinson’s disease^5–12^. Yet there are still few appropriate treatment options across these diseases, and diagnoses remain a challenge, as a direct result of a lack of full understanding regarding the neuropathologic mechanisms of the diseases^13–16^. To mitigate these gaps, greater effort must be made to combine resources in the pursuit of novel discovery in neurodegenerative disease genetics.

The application of open-science and data sharing principles in the pursuit of neurodegenerative disease research motivated the deployment of the Neurodegenerative Disease Knowledge Portal (NDKP)^17^, consisting of 218 open-access genomic summary statistics and variant datasets of individuals across ten different phenotypic groups, including neurological conditions such as Alzheimer’s disease, ALS, Lewy body dementia, and Parkinson’s disease (all sample sizes were obtained December 2023; annual updates to the NDKP are anticipated.). Here, we further describe the online, open-access resource including details of the available data and applications for genetic discovery and result replication in the study of neurodegenerative diseases.

### Overview of the Neurodegenerative Disease Knowledge Portal

To maximize the potential for discovery from the many large, novel datasets being leveraged for various neurodegenerative disease genetic association studies, and in taking inspiration from the successes of the previously deployed Type 2 Diabetes Knowledge Portal^18^, a centralized repository was assembled to securely store these datasets and make their summary results widely available to the research community. The portal was created and deployed by the developers at the Broad Institute of MIT and Harvard in collaboration with the Montreal Neurological Institute-Hospital, the Global Parkinson’s Genetics Program (GP2), and National Institutes of Health (NIH) Intramural Center for Alzheimer’s and Related Dementias (CARD) into what is now known as the NDKP.

Overall, the aim of the NDKP is to aggregate, enable analysis, and display all available genomic datasets of neurodegenerative disease, while protecting the integrity and confidentiality of the underlying datasets. The NDKP is available to the broad scientific community studying neurodegeneration seeking to unveil novel genetic associations or validate primary findings from other approaches.

### Available datasets

Although the goal of the NDKP is to expand our genomic understanding of neurodegenerative diseases, the portal includes genomic datasets from cohorts spanning ten phenotypic groups, including: 1) cerebrovascular magnetic resonance imaging (MRI) traits, 2) COVID-19, 3) immunological, 4) metabolite, 5) musculoskeletal, 6) neurological, 7) psychiatric, 8) sleep and circadian, 9) stroke, and 10) cognitive. Across these groups, 239 sub-phenotypes are captured (**eTable 1**). Importantly, this allows for the cross analysis of neurodegenerative diseases with possible related and overlapping features. For example, given the known association between cerebrovascular disease features and neurodegeneration risk^19–22^, the NDKP can be used to identify genes or single nucleotide polymorphisms (SNPs) relevant in both a neurological phenotype such as Alzheimer’s disease and a cerebrovascular MRI feature such as brain microbleeds, as will be discussed further below. In total, 218 genomic datasets are currently captured within the NDKP (**Figure 1**), which can be subdivided into two data types: 1) genotyping studies and 2) sequencing studies (**Figure 2**).

**Figure 1.**
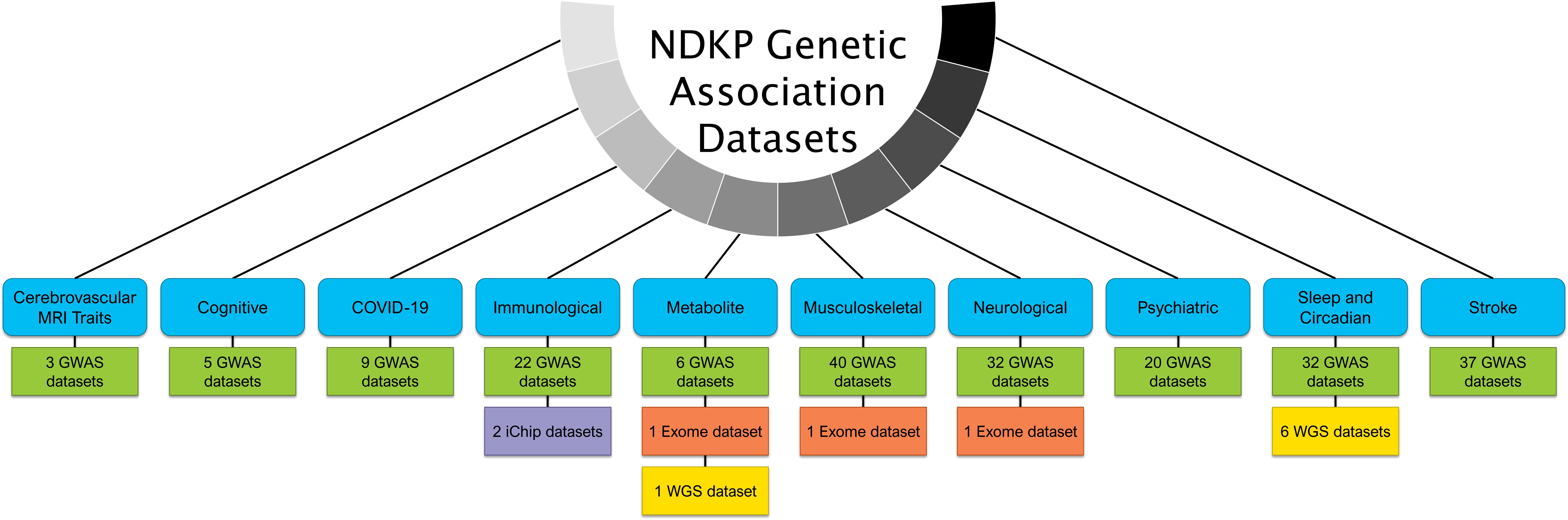
Genomic association datasets captured by the Neurodegenerative Disease Knowledge Portal (NDKP) from various phenotypic groups. The NDKP is comprised of genomic datasets from cohorts spanning nine phenotypic groups, including cerebrovascular magnetic resonance imaging (MRI) traits, COVID-19, immunological, metabolite, musculoskeletal, neurological, psychiatric, sleep and circadian, and stroke. Datasets include genotyping data, such as genome-wide association studies (GWAS) and ImmunoChip (iChip), and sequencing studies, such as whole genome sequencing (WGS) and exome sequencing.

**Figure 2.**
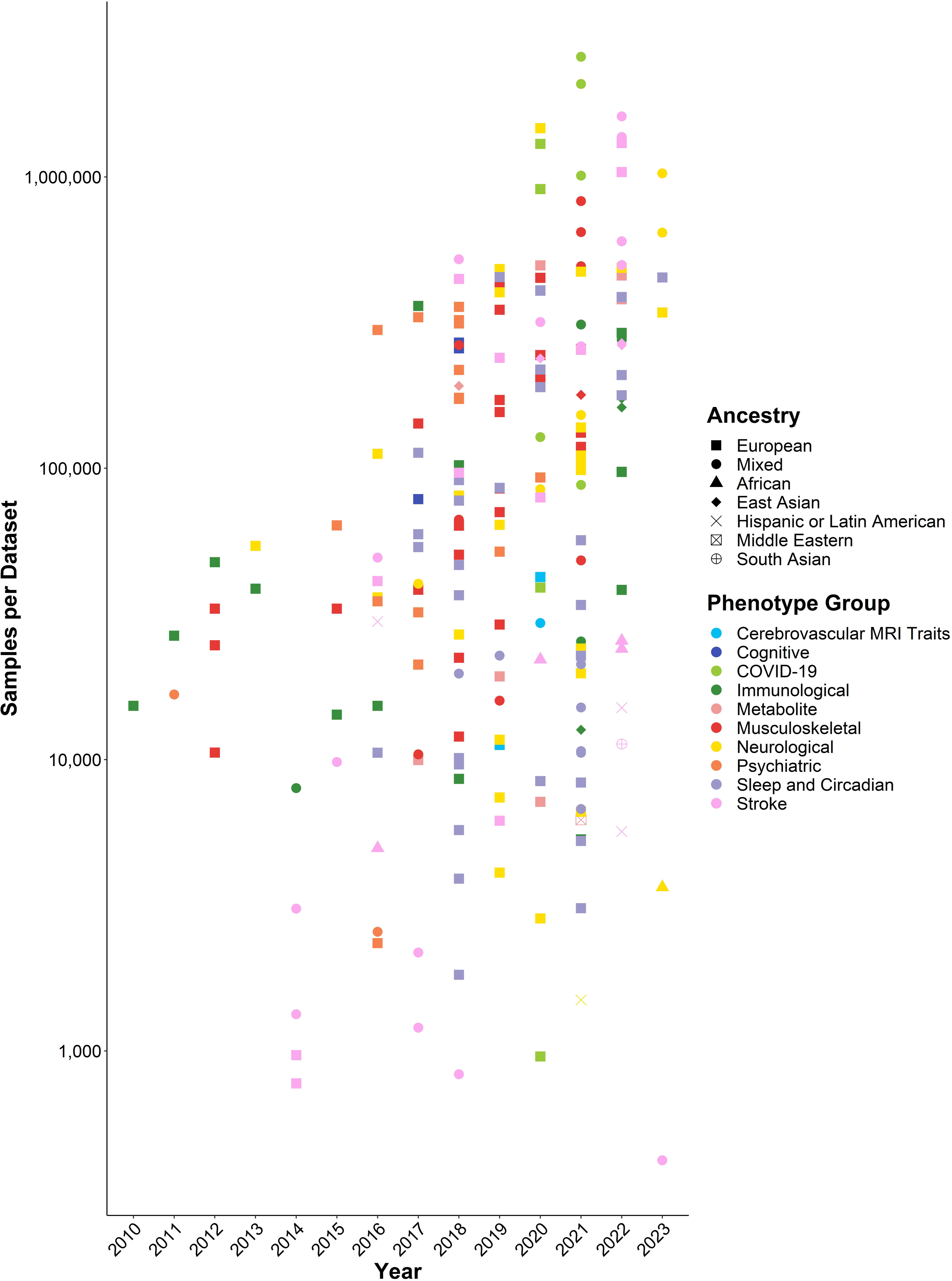
Year of publication and sample size of the 172 genomic association datasets captured by the Neurodegenerative Disease Knowledge Portal (NDKP) from various phenotypic groups. Datasets include genotyping data, such as genome-wide association studies (GWAS) and ImmunoChip (iChip), and sequencing studies, such as whole genome sequencing (WGS) and exome sequencing. For one study that provided pre-published data with no given date of publication, the year of publication 2023 was used. Samples per dataset are presented on a log10 scale.

#### Genotyping studies

The genotyping-based studies within the NDKP largely encompass genome-wide association studies (GWAS), which employ genotyping microarray data and are aimed at identifying associations between SNPs and traits or disease states. GWAS require large sample pools of both individuals with a phenotype of interest and non-affected controls, and depending on the type of quantitative trait, can generate an effect size per variant^23^. The NDKP includes datasets from 206 GWAS performed between 2010 and 2023, with the largest neurodegenerative disease dataset having been part of the 2023 GP2 GWAS that included 1,028,993 samples across multiple ancestries^24^ (**Figure 2**). Summary statistics from many of the GWAS, which include the aforementioned effect sizes per statistically significant, phenotype-associated SNP, are directly downloadable from the NDKP on the “Data Downloads” page.

In addition to the case-control variant calls and summary statistics, additional data types have been generated for the NDKP from the GWAS analyses. “Credible sets” include sets of variants near significant genetic association signals that are likely to include the causal variant for the signals and are generated through fine mapping of the GWAS results to further investigate genetic association signals. Additionally, “effector gene lists” encompass lists of variant-adjacent genes potentially mediating the effects of the significantly phenotype-associated variants identified within the GWAS. Although the methods by which these genes are defined are study-dependent, their inclusion in the NDKP is important as stepping stones for experiments to further define the genes that may be truly causal for a particular phenotype.

Finally, the NDKP also has two datasets from the immunological phenotype group that encompass ImmunoChip (iChip) data. The iChip is a custom-designed Illumina Infinium microarray that includes a specific set of SNPs and small insertion-deletions that were previously associated with autoimmune and inflammatory diseases through GWAS^25^. It provides the added benefit of being more cost-effective than typical exome- or genome-wide microarrays.

#### Sequencing studies

Sequencing studies include the data from whole genome, exome, or smaller panel-based next-generation sequencing (NGS) analyses. Typically, these studies include sequences from both affected (individuals with the phenotype of interest) and unaffected individuals, as the data are utilized to perform variant binned association analyses, such as rare variant burden association analysis. Generally, these approaches collapse rare variants into groups that can be dictated by a variety of factors, including, but not limited to: general genomic region, individual genes, pathways of interest, minor allele frequency, or functional consequence^26^.Various methods can be used to identify associations between the rare variant groups and the phenotype of interest, such as univariate or multivariate regression models. Importantly, sequencing data not only allows for analysis of variant-level associations but also for assessment of gene- and region-level associations, using variants called from the complete sequencing of all loci within a given region of the genome.

Currently, the NDKP includes the data from ten sequencing studies, including seven whole genome and three exome datasets (**Figure 2**). As NGS becomes more cost-effective, we expect larger and more ethnically diverse sequencing datasets to be made available and included in Knowledge Portal initiatives.

#### Neurological datasets

The NDKP includes 33 datasets within the neurological phenotype subgroup, including 32 GWAS datasets and one exome sequencing dataset (**Table 1**). These neurological phenotype datasets span eight sub-phenotypes, including Alzheimer’s disease (general and late-onset), ALS, Parkinson’s disease, epilepsy, and Lewy body dementia. While 26 of the datasets are of European ancestry, one dataset has subjects of African ancestry, another has subjects of Hispanic ancestry, and five datasets represent multi-ancestry analyses. In the coming years, it is a priority for the NDKP to continue expanding the ancestral diversity of the available datasets.

**Table 1.** Neurological datasets included in the Neurodegenerative Disease Knowledge Portal (NDKP)

| Dataset | Publication Year | Cases (n) | Controls (n) | Ancestry | Data Type |
| --- | --- | --- | --- | --- | --- |
| Alzheimer's disease GWAS | 2019 | 71880 | 383378 | European | GWAS |
| Alzheimer's disease GWAS | 2021 | 75024 | 397844 | European | GWAS |
| Alzheimer's disease GWAS | 2022 | 85934 | 401577 | European | GWAS |
| Alzheimer's disease GWAS | 2023 | 101061 | 543127 | Multi | GWAS |
| Alzheimer's disease family history GWAS | 2018 | 314278 | N/A* | European | GWAS |
| Amyotrophic lateral sclerosis exome case-control | 2019 | 3864 | 7839 | European | Exome |
| Amyotrophic lateral sclerosis GWAS | 2016 | 12577 | 23475 | European | GWAS |
| Amyotrophic lateral sclerosis GWAS | 2017 | 13811 | 26325 | Multi | GWAS |
| Amyotrophic lateral sclerosis GWAS | 2018 | 20806 | 59804 | European | GWAS |
| Amyotrophic lateral sclerosis GWAS | 2020 | 22040 | 62654 | Multi | GWAS |
| Amyotrophic lateral sclerosis GWAS | 2021 | 29612 | 122656 | Multi | GWAS |
| Amyotrophic lateral sclerosis GWAS | 2021 | 27205 | 110881 | European | GWAS |
| Carpal tunnel syndrome GWAS | 2019 | 12312 | 389334 | European | GWAS |
| Cognitive function GWAS | 2016 | 112067 | N/A | European | GWAS |
| FinnGen r8 complex disease GWAS (Alzheimer's disease) | 2023 | 7129 | 760059 | European | GWAS |
| Global Parkinson's Genetics Program GWAS | 2023 | 62976 | 966017 | Multi | GWAS |
| GR@CE Alzheimer's GWAS | 2019 | 4120 | 3289 | European | GWAS |
| Handedness GWAS | 2020 | 1470460 | N/A | European | GWAS |
| International League Against Epilepsy GWAS | 2018 | 225 | 24218 | European | GWAS |
| IPDGC Parkinson's disease GWAS (females) | 2021 | 7384 | 12389 | European | GWAS |
| IPDGC Parkinson's disease GWAS (males) | 2021 | 12054 | 11999 | European | GWAS |
| IPDGC-UK Biobank Parkinson's disease and proxy cases GWAS (female) | 2021 | 13420 | 90662 | European | GWAS |
| IPDGC-UK Biobank Parkinson's disease and proxy cases GWAS (male) | 2021 | 20956 | 89660 | European | GWAS |
| IPDGC-UK Biobank Parkinson's disease GWAS (female) | 2021 | 7947 | 90662 | European | GWAS |
| IPDGC-UK Biobank Parkinson's disease GWAS (male) | 2021 | 13020 | 89660 | European | GWAS |
| LARGE-PD Parkinson's disease GWAS | 2021 | 807 | 690 | Hispanic/Latin American | GWAS |
| Late-onset Alzheimer's GWAS | 2013 | 8572 | 11312 | European | GWAS |
| Late-onset Alzheimer's GWAS | 2019 | 21982 | 41944 | European | GWAS |
| Lewy body dementia GWAS | 2021 | 2981 | 4391 | European | GWAS |
| Parkinson's disease GWAS | 2019 | 56306 | 426424 | European | GWAS |
| Parkinson's disease GWAS | 2023 | 1200 | 2445 | African | GWAS |
| Parkinson's disease progression GWAS | 2019 | 4093 | N/A | European | GWAS |
| Parkinson's disease progression GWAS | 2020 | 2848 | N/A | European | GWAS |

Further details regarding the neurological datasets captured by the Neurodegenerative Disease Knowledge Portal can be accessed online at: https://ndkp.hugeamp.org/datasets.html
\*25,696 individuals had a maternal family history of Alzheimer's disease and 14,338 individuals had a paternal family history of Alzheimer's disease. Abbreviations: GR@CE, Genome Research at Fundacio ACE; GWAS, genome-wide association study; N/A, not applicable.

#### Multi-omic datasets

In addition to the genomic datasets available through the NDKP, multi-omic data from the Common Metabolic Diseases Genome Atlas (CMDGA) are integrated through the various search pages and tools provided by the portal^18^, as discussed further below. Briefly, the CMDGA provides a compendium of epigenomic and other functional genomic data collated from the Accelerating Medicines Partnership Common Metabolic Disorders (AMP-CMD) consortium, publicly available sources, and large resources such as ChIP-Atlas and the Encyclopedia of DNA Elements (ENCODE). In total, there are 6890 multi-omic datasets encompassed within the NDKP that were produced using a wide-variety of methods including, but not limited to, ATAC-Seq, CaptureC, ChIP-Seq, HiC, and RNA-Seq. Of these, 378 tissues are derived from tissues relevant to the central nervous system. Importantly, these datasets allow for the annotation of variants captured within the NDKP based on whether they are encompassed by regions considered accessible chromatin, binding sites, candidate regulatory elements, chromatin state, gene expression, histone modifications, or target variant predictions.

### Bioinformatics processing of the data

Upon intake, genetic and genomic datasets are subjected to a suite of bioinformatic methods to glean additional insights from the processed and integrated results. Incoming genetic association datasets are first subjected to quality control and harmonization, including ensuring that standardized column headings are utilized, inferring missing data for non-optional columns (e.g. odds ratios can be used to infer effect sizes), and lifting over all datasets to GRCh37. We also ensure that all effect sizes are in reference to the alternate allele of GRCh37, remove variants with incompatible summary statistics for the subsequent analyses, and perform a linear regression-based effect size scaling for all quantitative phenotypes^18^. Variant associations are then meta-analyzed, using the METAL algorithm that infers and accounts for sample overlap between datasets to calculate an integrated “bottom-line” association for each variant and each trait^27^. The bottom-line analysis can both identify novel associations that are not significant at the level of individual datasets but become significant when multiple studies are considered and identify artifactual associations that may be significant in one dataset but are not replicated in others using a fixed effect method. Using these bottom-line associations, we run the Variant Effect Predictor^28^ to annotate predicted variant impact and perform LD-clumping using the PLINK method^29^, to group variants into genetically linked sets. We use the MAGMA algorithm per gene^30^, to calculate gene-level association scores based on nearby common variant associations, and per trait, to generate lists of biological pathways whose constituent genes are enriched for genetic associations for that trait. We apply the LD score regression method (LDSC) in two calculations^31, 32^. Cross-trait LDSC is used to calculate the genetic correlations between all traits, while stratified LDSC provides a measure of the enrichment of genetic association signals for each trait within annotated genomic regions such as enhancers and promoters. Finally, we apply the Human Genetic Evidence Calculator (HuGE Calculator) across all associations to categorize the weight of evidence supporting the relevance of each gene to each trait^33^. These methods are documented in the “Help” pages of the NDKP and were also described in detail by Costanzo et al.^18^.

### Application for the study of neurodegeneration

In addition to securely hosting large genomic datasets, one of the founding aims of the NDKP is to provide accessible workflows and tools to effectively utilize the datasets and assist in the facilitation of customized genomic analyses. To allow users to carry out these aims, the portal offers four core page types, including regional pages, gene pages, variant pages, and phenotype pages, in addition to a variety of tools to allow for more structured analyses. Across these pages and tools, users are provided with summary results derived from the genomic datasets to explore genes, genomic regions, variants, or phenotypes of interest (**Figure 3**).

**Figure 3.**
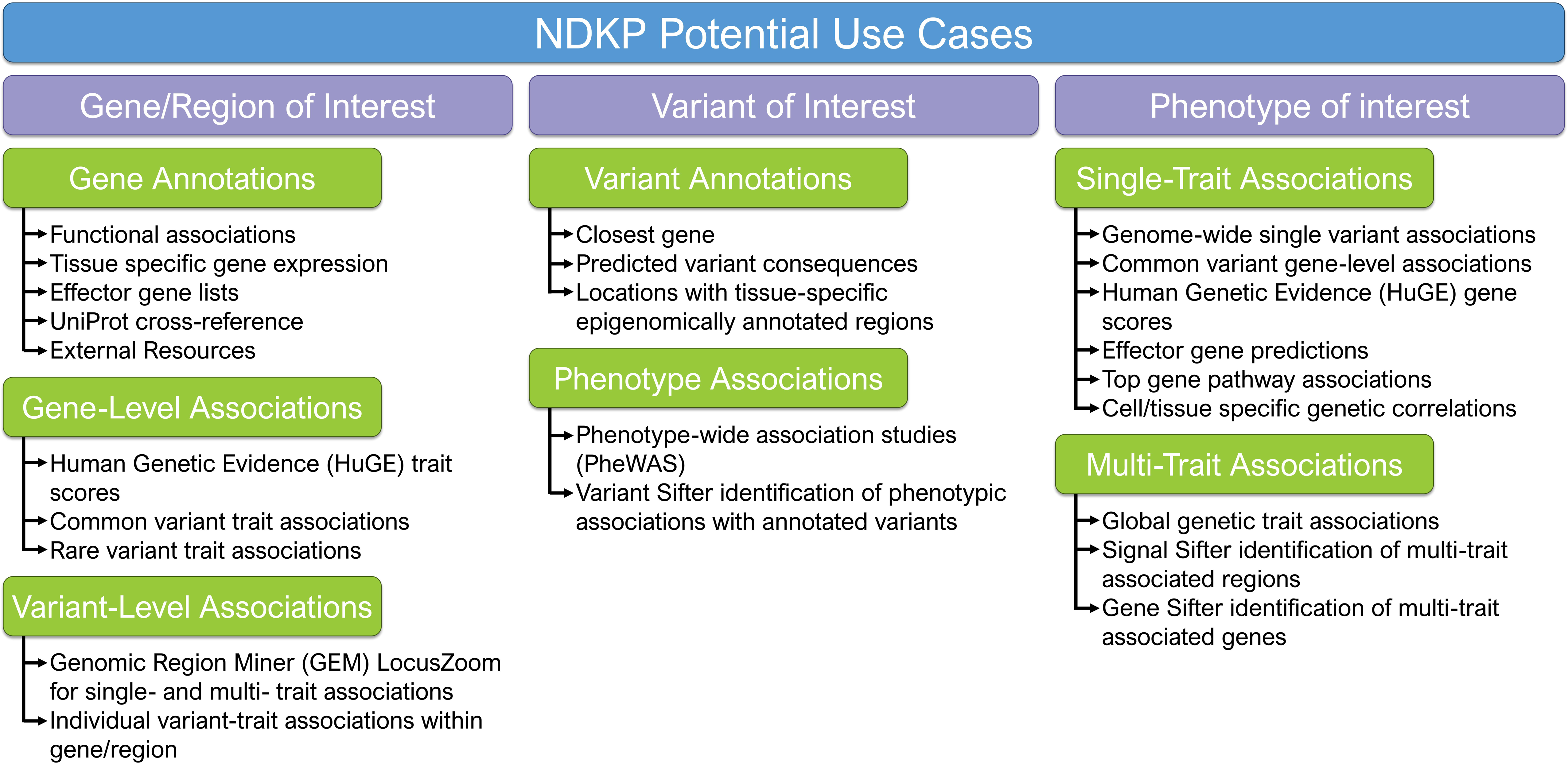
Potential uses for the data and tools encompassed within the Neurodegenerative Disease Knowledge Portal (NDKP). The NDKP aims to provide accessible workflows and tools to utilize the datasets and assist in the facilitation of customized genomic analyses. The Portal offers four core search pages and a variety of tools to provide summary results derived from the genomic datasets to explore genes, genomic regions, variants, or phenotypes of interest.

#### Exploring genes/regions of interest

Commonly, researchers from disciplines outside the realm of computational or statistical genomics identify genes or regions of the genome of interest from experimental study, such as comparative functional genomic analyses with model organisms, protein-protein interaction analyses, or expression quantitative trait loci (eQTL) analyses^34, 35^. Following such studies, researchers may wish to further explore whether human genetic and genomic results support the hypotheses generated via experimentation. However, it has often been difficult for these research groups to gain access to the necessary large-scale genotyping or sequencing datasets to explore these results. Further, even if data were freely available, these researchers may have lacked the expertise to efficiently or accurately pull the required summary-level results to substantiate their hypotheses. The NDKP has aimed to fill this gap, providing a variety of results that may be of interest both to the genetics community and to researchers outside of the discipline to explore regions of the genome and specific genes that may represent new risk loci or therapeutic targets for specific neurodegenerative phenotypes (**Figure 3**).

Following the search of a gene of interest, the NDKP returns a variety of gene- and variant-level association summary results. At the gene-level, the search returns both common variant and rare variant gene-wide trait associations. More specifically, these results represent phenotypes for which a genetic association exists with the genes when common or rare variants, respectively, are binned and the burden of variants within the gene are compared between a cohort of individuals with the phenotype to a control cohort that does not have the phenotype. The NDKP also returns HuGE Calculator trait scores for the gene, which are calculated from a method created by Dornbos et al. and represent the extent of human genetic evidence captured within the Knowledge Portals that supports gene-phenotype associations^33^. For example, the well-established ALS-associated gene, *SOD1*, has a calculated HuGE score of 350, representing a “compelling” level of evidence for involvement of the gene in ALS, whereas the gene *LDLR*, which is traditionally associated with familial hypercholesterolemia and has no known associations with ALS, has a calculated HuGE score of 1.33, representing “anecdotal” evidence.

In addition to gene-level summary results, the search of a gene in the NDKP will also return variant-level summaries, including a list of variants within the gene that have individual associations with any of the phenotypes captured across the various datasets. Similarly, the search of a region of interest will return individual variant-level trait associations that have been identified across the NDKP datasets within the given region. The search of a region of interest will also return the Genomic Region Miner (GEM) LocusZoom tool, which visualizes variant associations with single or multiple traits. **Figure 4** displays the results of searching for the region surrounding a well-known Parkinson’s disease gene, *SNCA* (chr 4:90,571,496-90,809,466). Using the GEM LocusZoom tool, not only can the variants with significant Parkinson’s disease associations be visualized, but if other trait-associations are suspected in the region, as is the case for Lewy body dementia in this region, additional phenotypes can be queried. A table of the variant-level data captured by the GEM LocusZoom tool is also provided by the NDKP.

**Figure 4.**
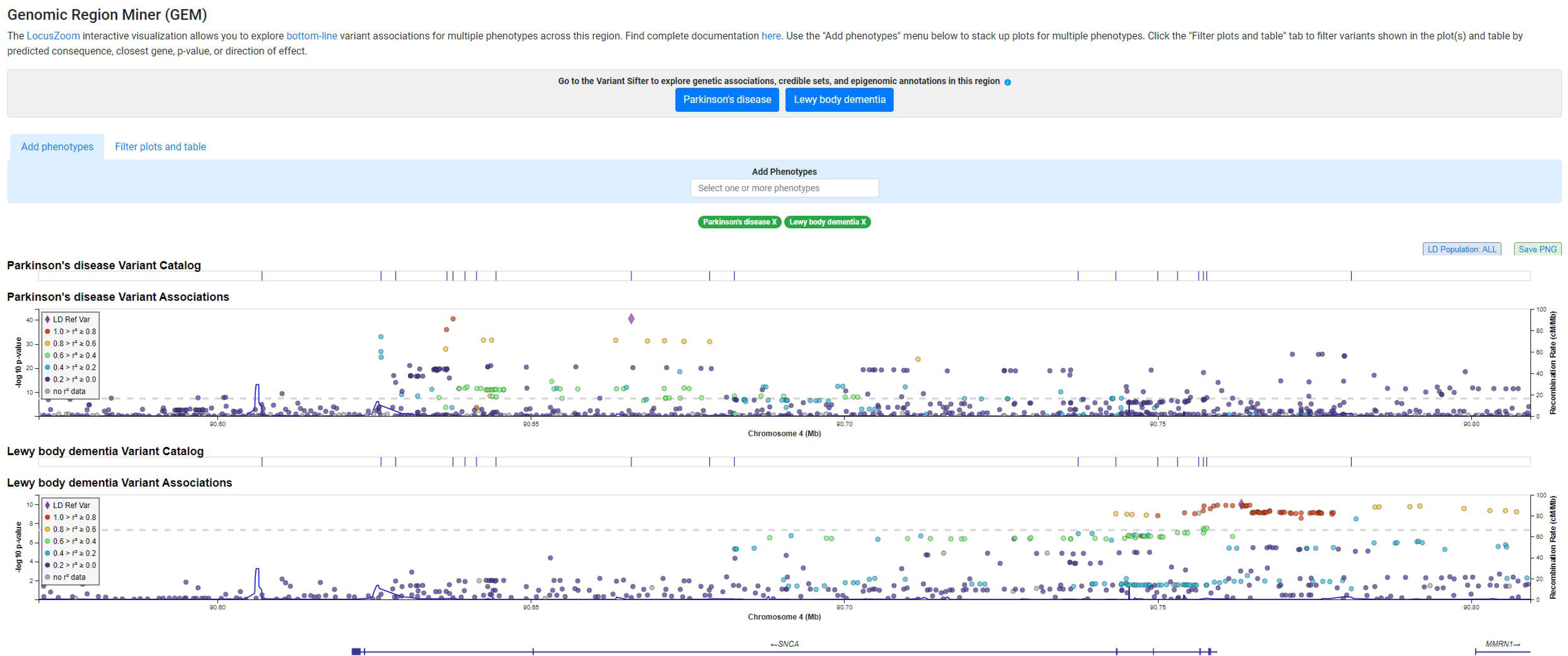
Genomic Region Miner (GEM) LocusZoom visualization of variant-phenotype associations within the region surrounding *SNCA* (chr 4:90,571,496-90,809,466). As part of the summary results returned when a region of interest is searched in the Neurodegenerative Disease Knowledge Portal (NDKP), the GEM LocusZoom tool provides a visualization of all variants within that region identified across the NDKP datasets and their associations with the regions most highly associated phenotype. In the case of the region surrounding *SNCA*, Parkinson’s disease represents the most highly associated phenotype. However, the GEM LocusZoom tool also allows for customized visualization of variant associations with additional phenotypes, as is shown here for Lewy body dementia.

In addition to the genetic association summary results computed and returned by the NDKP, a variety of gene annotations can be found from the search of a given gene or region. The search of a gene of interest returns functional associations of the gene, tissue specific gene expression data, effector gene lists, UniProt cross-reference data, and external resource links as applicable. In contrast, the search of a region of interest returns a list of genes encompassed by the region.

#### Exploring variants of interest

Variants of interest, such as those identified through GWAS analysis or other association studies, can also be further explored using the NDKP. The search of a variant using the dbSNP identifier will return both variant annotations and phenotype associations based on the datasets encompassed in the portal (**Figure 3**). More specifically, the variant search page will return information regarding the closest gene to the variant or the gene it resides within, as applicable, as well as any predicted variant consequences. The search page also returns results of a phenotype-wide association study (PheWAS), providing the statistical results that describe the level of associations of the variant with any given phenotype captured by the NDKP. For example, when a search of the NDKP is performed for the *APOE* e4 defining SNP, rs429358, an unsurprising significant association is observed between the variant and an increased risk of late-onset Alzheimer’s disease (OR = 3.49, p = 4.94e-324); however, the variant is also found to be significantly associated with an increased risk of brain microbleeds (OR = 1.29, p = 7.48e-10). Importantly, individual datasets can also be specifically queried for variants of interest, which can provide interesting sources of evidence for ancestry specific analyses or variant curation exercises^36^.

#### Exploring phenotypes of interest

Unlike the above examples that are often driven by experimentally derived hypotheses, a researcher may also have a phenotype or disease of particular interest for which they want to develop novel human genomic derived hypotheses. For these instances, the NDKP provides phenotype search page result summaries in addition to specific tools that allow for the exploration of genetic associations (**Figure 3**).

When only a single phenotype is of interest, the phenotype search page provides the greatest amount of information to the user, including both variant-level and gene-level result summaries. All datasets for which the phenotype of interest is captured are clearly outlined. At the variant level, the phenotype search page provides the top genome-wide single-variant associations, and at the gene-level, associations based on the binning of common variants within each potential gene are provided. The search also provides top gene pathway associations, cell/tissue specific genetic correlations, and effector gene predictions for the phenotype, as applicable.

Using ALS as a case study, the user will find seven datasets that capture the ALS phenotype, each of which can be further explored. As anticipated, the top single-variant association signal for ALS is an intronic SNP (rs2453555) within *C9orf72* (OR = 1.19, p = 1.78e-41), which tags a hexanucleotide repeat expansion known to be one of the most common genetic causes of ALS^37–40^. Similarly, *C9orf72* (p = 3.04e-20, variants = 24) is the gene with the second highest common variant gene-level association, following *MOB3B* (p = 6.53e-29, variants = 95), a gene located nearby *C9orf72*. The NDKP also indicates that ALS is significantly associated with the acanthocytosis pathway (p = 3.22e-7) — an example of a potential novel association that may prompt hypothesis generation — and ALS genetic associations are significantly enriched within regions annotated as enhancers in central nervous system tissues (p = 7.07e-5). In addition to the results captured by the phenotype search pages, the NDKP also assists with single-trait analysis through the use of the HuGE Calculator that computes HuGE scores for any given phenotype-gene combination, which are described further above^33^.

Of additional utility, the NDKP also offers summary results and tools that allow for effective multi-trait analysis. Directly within the phenotype search page, a list of genetically correlated traits for the phenotype of interest can be found. Using Alzheimer’s disease as a case study, it is unsurprising to find a significant genetic correlation with late-onset Alzheimer’s disease (r = 0.90, p = 1.22e-70); however, there is also a significant genetic correlation observed with Parkinson’s disease (r = 0.26, p = 7.80e-8).

In addition, the NDKP hosts three tools that allow for more detailed multi-trait analysis: 1) the Signal Sifter, 2) the Gene Sifter, and 3) the Variant Sifter. The Signal Sifter and Gene Sifter tools work in a similar manner, such that multiple traits can be queried and genetic associations relevant to two or more phenotypes will be returned, but the Signal Sifter returns regions with LD-clumped variants, while the Gene Sifter returns genes. Often the user will explore genetic correlations by beginning with multiple phenotypes that they know, or suspect, are clinically correlated. Importantly, the integration of these tools directly within the framework of the NDKP affords researchers the ability to explore the genetic underpinnings of common comorbidities across the neurodegenerative disease spectrum.

Again, using Alzheimer’s disease as a primary phenotype, the user may wish to explore its genetic correlations with brain microbleeds based on observations of neurovascular damage that have been observed in cases of neurodegenerative disease^41–43^. Indeed, upon investigation of these two phenotypes with the NDKP, the Signal Sifter returns regions of LD-clumped variants significantly associated with increased risk of both traits, including the top associated region chr 19:45,387,459-45,428,235 (Alzheimer’s associated variant rs11556505, p = 4.94e-324; brain microbleed associated variant rs769449, p = 2.52e-10) (**Figure 5A**). Similarly, the Gene Sifter returns 138 genes with significant chi-square p-values that represent a measure of overall association for the gene and both traits (**Figure 5B**). The top three of these associated genes are *TOMM40* (p[X^2^] = 1.14e-141), *PVRL2* (p[X^2^] = 7.40e-137), and *APOE* (p[X^2^] = 2.68e-102), expectedly, all of which are captured in the top region identified with Signal Sifter and have been previously associated with the two phenotypes independently^10, 44–48^.

**Figure 5.**
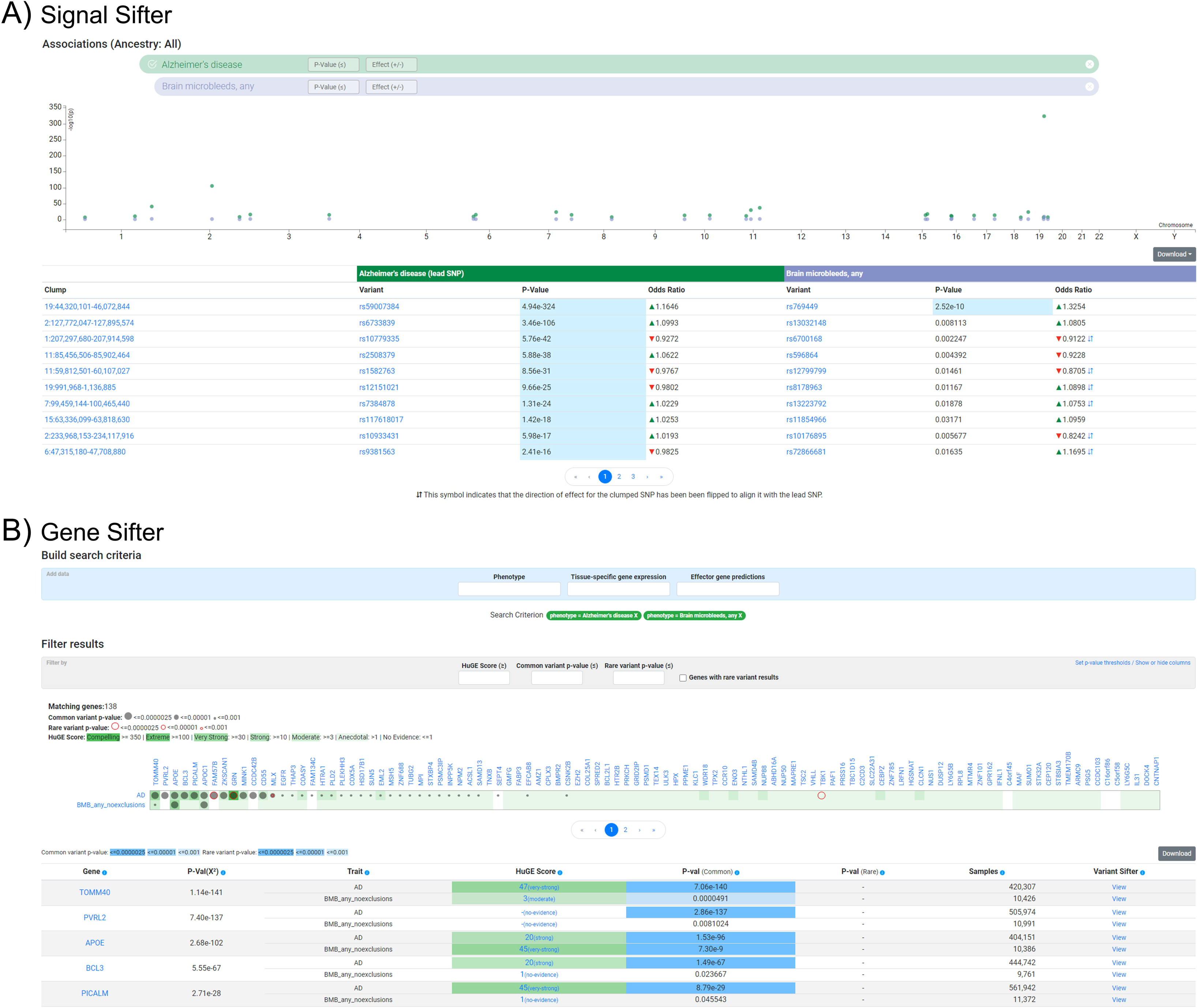
Multi-trait analysis using tools integrated into the NDKP, which demonstrated genomic associations with Alzheimer’s disease and brain microbleeds. **(A)** Signal Sifter identified regions of LD-clumped variants significantly associated with risk of both traits. **(B)** Gene Sifter identified 138 genes with a significant chi-square p-value, indicating overall associations between the genes and both traits.

The Variant Sifter is the Knowledge Portal’s most recently developed tool and encompasses a wide range of capabilities, broadly allowing the user to explore variant-phenotype associations based on a range of filter options including focusing on credible sets and tissue-specific epigenomic annotations. Although there are many ways to utilize the tool, the user typically will begin with a phenotype and a region or gene of interest. The Variant Sifter returns a list of variants in the region that are associated with the phenotype that can then be filtered based on user-defined criteria. The filters allow for the identification of variants in credible sets, variants within tissue-specific regulatory region annotations of interest — integrated from the CMDGA — and variants linked to specific genes.

Recently, a novel ALS-associated gene, *KANK1*, was discovered using a rare variant association analysis approach, which not only identified an enrichment of rare variants in coding regions of the gene in individuals with ALS but also rare variants in non-coding enhancer and promoter regions^7^. The NDKP Variant Sifter can be used to determine if any individual variants within the surrounding regulatory regions of *KANK1* demonstrate a significant association with ALS, which may be important for further functional analyses regarding this gene’s association. Following the search of the chr9:370,291-846,105 region using the Variant Sifter tool with respect to the ALS phenotype, the tool returns all variants reported within the NDKP in an association plot (**Figure 6A**). The variants can then be filtered based on a variety of annotations of interest to the researcher, such as identifying variants by location within regulatory regions annotated in broad tissue categories based on the epigenomic data derived from the CMDGA. In this case, we were interested in identifying variants within an enhancer regulatory element in the central nervous system linked to *KANK1*. We first filtered for variants located within enhancer regions and specified to only include those identified in tissues of the central nervous tissue (**Figure 6B**). The Variant Sifter then provides a filter to identify variants linked to genes, using which we specified to only include variants linked to *KANK1* (**Figure 6C**). Using this filtration strategy that leveraged the epigenomic data derived from the CMDGA, the Variant Sifter tool returned 79 variants, including one variant (chr9:504,491:A>C) demonstrating an association with ALS based on the meta-analysis of NDKP data that is approaching significance (β = 0.238, p = 7.01e-4; **Figure 6D**).

**Figure 6.**
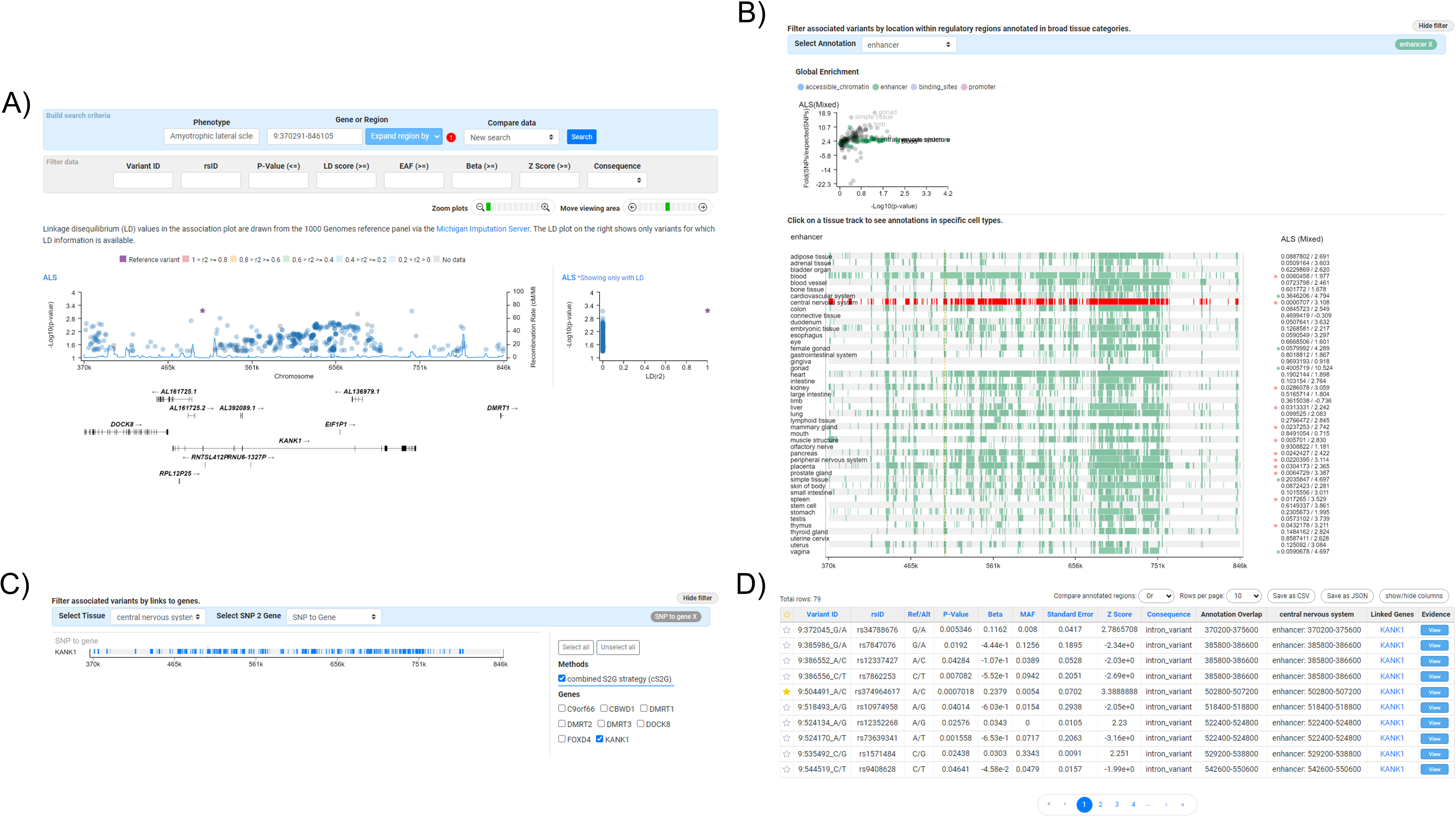
Exploration and filtration of variant associations with amyotrophic lateral sclerosis (ALS) within the surrounding region of *KANK1* (chr9:370291-846105). **A)** Association plot of all variants identified within the meta-analyzed datasets encompassed within the Neurodegenerative Disease Knowledge Portal (NDKP) within the surrounding region of *KANK1* in reference to their meta-analyzed associations with ALS. **B)** Filtration of variants to only those in enhancer regulatory element within the central nervous system based on epigenomics data encompassed within the NDKP from the Common Metabolic Diseases Genome Atlas (CMDGA). **C)** Filtration of variants to include only those linked to *KANK1*. **D)** All remaining variants following the annotation filters applied using the Variant Sifter tool. The star represents a variant of interest that has been manually selected based on its association with ALS that is approaching significance.

## Discussion

As researchers and consortia continue to expend immense effort and resources in producing large-scale genomic analyses, it is essential that the greatest potential for discovery is realized from these datasets. The NDKP offers a centralized knowledge base for researchers that can act as a secure, accessible, and innovative solution for data sharing, not only achieving aims of transparency when reporting novel results, but also allowing for continued discovery in neurodegenerative disease research. In contrast to disease-agnostic resources that aggregate large-scale genomic datasets, such as Open Targets^49^ or the GWAS Catalog^50^, the NDKP focuses the presented results on traits most relevant to neurodegenerative phenotypes and benefits from the careful curation provided by neurogenetic experts to ensure all analyses of high impact are incorporated appropriately and displayed in ways most useful to the end-user. Yet, unlike highly disease-specific “omic” resources, such as the Alzheimer’s Disease Variant Portal^51^ or Agora^52^, the NDKP still allows for the assessment of co-morbidities across the neurodegenerative spectrum, which is particularly important based on the large amount of clinical overlap between these conditions. Furthermore, the various user interfaces and integration of analytic tools are custom-designed for the purposes of the Knowledge Portals and enable consistent extraction of high-quality results at the source, for both genomics experts and those in other disciplines, while still protecting the integrity of the data.

In addition to its potential for discovery, the NDKP provides a centralized hub allowing the user to replicate or further investigate findings from their own independent datasets. The generation of “omics” datasets can be cost prohibitive, particularly considering the need for both discovery and replication subsets^53^; by offering a secondary dataset for replication of novel discoveries, researchers can maximize the statistical power harnessed from their in-house data. Additionally, the breadth of data types and tools available through the portal offer the ability to further explore their findings beyond only replication of results. Our goal is to continue to add available datasets to the portal. Regular data releases are planned, at minimum on a yearly basis, including the addition of at least nine new datasets spanning multiple neurodegenerative diseases and diverse ancestral populations. We aim to prioritize the integration of additional “omic” datatypes, such as single-cell, bulk RNA-seq, and proteomics analyses, in addition to incorporating deeply phenotyped clinical datasets as they become available. As we move towards achieving this goal, we also remain committed to the development of appropriate data mining methods and bioinformatics tools to appropriately utilize the data, which is particularly important for complex phenotypes such as neurodegenerative diseases^54^. Finally, we welcome new collaborations, including the opportunity to incorporate additional data, methods, and tools into the NDKP. Researchers are encouraged to contact the data intake team to discuss prospective collaborations and data deposition.

Although effort is being made to expand the portal, it is important to recognize that the NDKP currently has inherent limitations regarding the available data. Most notably, there is an overrepresentation of individuals of European ancestry, largely reflecting the lack of diversity observed across the field of genomics^55^. Further, most currently available datasets represent GWAS summary statistics, which do not represent all information from the original genotyping microarray data. While offering potential for further discovery and replication of common variant signals, GWAS datasets typically do not capture rare genetic biomarkers for phenotypes of interest.

Here, we have provided a comprehensive overview of the vast amount of data included in the NDKP and their possible applications, yet there are unlimited potential use cases. Additional detailed potential workflows have been outlined by the Knowledge Portal developers, including using the NDKP to perform multi-trait analysis, rare variant association gene-level analysis, and integrative analysis available within the “Workflow” subsection of the “Help” pages. The developers have also generated ample tutorials, webinars, and presentations to aid in NDKP use available within the “Videos, webinars, and presentations” subsection of the “Help” pages. Ultimately, our goal is for available data to be accessible and easy to use for both novel discovery and replication purposes, promoting open-science and collaboration, and resulting in reproducible conclusions that will improve target discovery for neurodegenerative diseases.

## Supporting information

eTable 1

## Data Availability

All data produced are available online in The Neurodegenerative Disease Knowledge Portal (NDKP; https://ndkp.hugeamp.org/)

## Declaration of interests

H.L.L., D.V., H.I., and M.A.N.’s participation in this project was part of a competitive contract awarded to Data Tecnica International LLC by the National Institutes of Health to support open science research. M.A.N. also currently serves on the scientific advisory board for Clover Therapeutics and is a scientific founder at Neuron23 Inc, he also owns stocks.

## Funding

This research was supported in part by the Intramural Research Program of the NIH, National Institute on Aging (NIA), National Institutes of Health, Department of Health and Human Services; project number ZO1 AG000534, as well as the National Institute of Neurological Disorders and Stroke. It was also supported by the Global Parkinson’s Genetics Program (GP2). GP2 is funded by the Aligning Science Across Parkinson’s (ASAP) initiative and implemented by The Michael J. Fox Foundation for Parkinson’s Research (https://gp2.org). For a complete list of GP2 members see https://gp2.org.

## Notes

### Author Declarations

The study used only openly available human data located within The Neurodegenerative Disease Knowledge Portal (NDKP; https://ndkp.hugeamp.org/)

### Summary of Updates

Minor revisions to the Available Datasets, Application for the Study of Neurodegeneration, and Discussion sections. Correction of an authorship error.

## References

1. Bocchetta M, Mega A, Bernardi L, et al. Genetic Counseling and Testing for Alzheimer’s Disease and Frontotemporal Lobar Degeneration: An Italian Consensus Protocol. J Alzheimers Dis 2016;51:277–291.

2. Pang SY, Teo KC, Hsu JS, et al. The role of gene variants in the pathogenesis of neurodegenerative disorders as revealed by next generation sequencing studies: a review. Transl Neurodegener 2017;6:27.

3. Postuma RB, Berg D, Stern M, et al. MDS clinical diagnostic criteria for Parkinson’s disease. Mov Disord 2015;30:1591–1601.

4. Strong MJ, Abrahams S, Goldstein LH, et al. Amyotrophic lateral sclerosis - frontotemporal spectrum disorder (ALS-FTSD): Revised diagnostic criteria. Amyotroph Lateral Scler Frontotemporal Degener 2017;18:153–174.

5. Farhan SMK, Howrigan DP, Abbott LE, et al. Exome sequencing in amyotrophic lateral sclerosis implicates a novel gene, DNAJC7, encoding a heat-shock protein. Nat Neurosci 2019;22:1966–1974.

6. Nicolas A, Kenna KP, Renton AE, et al. Genome-wide Analyses Identify KIF5A as a Novel ALS Gene. Neuron 2018;97:1268–1283 e1266.

7. Zhang S, Cooper-Knock J, Weimer AK, et al. Genome-wide identification of the genetic basis of amyotrophic lateral sclerosis. Neuron 2022;110:992–1008 e1011.

8. Chang D, Nalls MA, Hallgrimsdottir IB, et al. A meta-analysis of genome-wide association studies identifies 17 new Parkinson’s disease risk loci. Nat Genet 2017;49:1511–1516.

9. Makarious MB, Lake J, Pitz V, et al. Large-scale rare variant burden testing in Parkinson’s disease. Brain 2023;146:4622–4632.

10. Bertram L, McQueen MB, Mullin K, Blacker D, Tanzi RE. Systematic meta-analyses of Alzheimer disease genetic association studies: the AlzGene database. Nat Genet 2007;39:17–23.

11. Bellenguez C, Kucukali F, Jansen IE, et al. New insights into the genetic etiology of Alzheimer’s disease and related dementias. Nat Genet 2022;54:412–436.

12. Prokopenko D, Lee S, Hecker J, et al. Region-based analysis of rare genomic variants in whole-genome sequencing datasets reveal two novel Alzheimer’s disease-associated genes: DTNB and DLG2. Mol Psychiatry 2022;27:1963–1969.

13. Duraes F, Pinto M, Sousa E. Old Drugs as New Treatments for Neurodegenerative Diseases. Pharmaceuticals (Basel) 2018;11.

14. Beber BC, Chaves MLF. Evaluation of patients with behavioral and cognitive complaints: misdiagnosis in frontotemporal dementia and Alzheimer’s disease. Dement Neuropsychol 2013;7:60–65.

15. Bicchi I, Emiliani C, Vescovi A, Martino S. The Big Bluff of Amyotrophic Lateral Sclerosis Diagnosis: The Role of Neurodegenerative Disease Mimics. Neurodegener Dis 2015;15:313–321.

16. Selvackadunco S, Langford K, Shah Z, et al. Comparison of clinical and neuropathological diagnoses of neurodegenerative diseases in two centres from the Brains for Dementia Research (BDR) cohort. J Neural Transm (Vienna) 2019;126:327–337.

17. The Neurodegenerative Disease Knowledge Portal [online]. Available at: https://ndkp.hugeamp.org/. Accessed December 10, 2023.

18. Costanzo MC, von Grotthuss M, Massung J, et al. The Type 2 Diabetes Knowledge Portal: An open access genetic resource dedicated to type 2 diabetes and related traits. Cell Metab 2023;35:695–710 e696.

19. Schneider JA, Arvanitakis Z, Bang W, Bennett DA. Mixed brain pathologies account for most dementia cases in community-dwelling older persons. Neurology 2007;69:2197–2204.

20. Seidel GA, Giovannetti T, Libon DJ. Cerebrovascular disease and cognition in older adults. Curr Top Behav Neurosci 2012;10:213–241.

21. Kummer BR, Diaz I, Wu X, et al. Associations between cerebrovascular risk factors and parkinson disease. Ann Neurol 2019;86:572–581.

22. Lendahl U, Nilsson P, Betsholtz C. Emerging links between cerebrovascular and neurodegenerative diseases-a special role for pericytes. EMBO Rep 2019;20:e48070.

23. Bush WS, Moore JH. Chapter 11: Genome-wide association studies. PLoS Comput Biol 2012;8:e1002822.

24. Kim JJ, Vitale D, Otani DV, et al. Multi-ancestry genome-wide association meta-analysis of Parkinson’s disease. Nat Genet 2024;56:27–36.

25. Cortes A, Brown MA. Promise and pitfalls of the Immunochip. Arthritis Res Ther 2011;13:101.

26. Asimit J, Zeggini E. Rare variant association analysis methods for complex traits. Annu Rev Genet 2010;44:293–308.

27. Willer CJ, Li Y, Abecasis GR. METAL: fast and efficient meta-analysis of genomewide association scans. Bioinformatics 2010;26:2190–2191.

28. McLaren W, Gil L, Hunt SE, et al. The Ensembl Variant Effect Predictor. Genome Biol 2016;17:122.

29. Purcell S, Neale B, Todd-Brown K, et al. PLINK: a tool set for whole-genome association and population-based linkage analyses. Am J Hum Genet 2007;81:559–575.

30. de Leeuw CA, Mooij JM, Heskes T, Posthuma D. MAGMA: generalized gene-set analysis of GWAS data. PLoS Comput Biol 2015;11:e1004219.

31. Finucane HK, Bulik-Sullivan B, Gusev A, et al. Partitioning heritability by functional annotation using genome-wide association summary statistics. Nat Genet 2015;47:1228–1235.

32. Bulik-Sullivan B, Finucane HK, Anttila V, et al. An atlas of genetic correlations across human diseases and traits. Nat Genet 2015;47:1236–1241.

33. Dornbos P, Singh P, Jang DK, et al. Evaluating human genetic support for hypothesized metabolic disease genes. Cell Metab 2022;34:661–666.

34. Zhu M, Zhao S. Candidate gene identification approach: progress and challenges. Int J Biol Sci 2007;3:420–427.

35. Nica AC, Dermitzakis ET. Expression quantitative trait loci: present and future. Philos Trans R Soc Lond B Biol Sci 2013;368:20120362.

36. Richards S, Aziz N, Bale S, et al. Standards and guidelines for the interpretation of sequence variants: a joint consensus recommendation of the American College of Medical Genetics and Genomics and the Association for Molecular Pathology. Genet Med 2015;17:405–424.

37. Byrne S, Elamin M, Bede P, et al. Cognitive and clinical characteristics of patients with amyotrophic lateral sclerosis carrying a C9orf72 repeat expansion: a population-based cohort study. Lancet Neurol 2012;11:232–240.

38. Majounie E, Renton AE, Mok K, et al. Frequency of the C9orf72 hexanucleotide repeat expansion in patients with amyotrophic lateral sclerosis and frontotemporal dementia: a cross-sectional study. Lancet Neurol 2012;11:323–330.

39. Umoh ME, Fournier C, Li Y, et al. Comparative analysis of C9orf72 and sporadic disease in an ALS clinic population. Neurology 2016;87:1024–1030.

40. Mejzini R, Flynn LL, Pitout IL, Fletcher S, Wilton SD, Akkari PA. ALS Genetics, Mechanisms, and Therapeutics: Where Are We Now? Front Neurosci 2019;13:1310.

41. Ramirez J, Berezuk C, McNeely AA, Scott CJ, Gao F, Black SE. Visible Virchow-Robin spaces on magnetic resonance imaging of Alzheimer’s disease patients and normal elderly from the Sunnybrook Dementia Study. J Alzheimers Dis 2015;43:415–424.

42. Raz L, Knoefel J, Bhaskar K. The neuropathology and cerebrovascular mechanisms of dementia. J Cereb Blood Flow Metab 2016;36:172–186.

43. Akoudad S, Wolters FJ, Viswanathan A, et al. Association of Cerebral Microbleeds With Cognitive Decline and Dementia. JAMA Neurol 2016;73:934–943.

44. Lambert JC, Ibrahim-Verbaas CA, Harold D, et al. Meta-analysis of 74,046 individuals identifies 11 new susceptibility loci for Alzheimer’s disease. Nat Genet 2013;45:1452–1458.

45. Lyall DM, Munoz Maniega S, Harris SE, et al. APOE/TOMM40 genetic loci, white matter hyperintensities, and cerebral microbleeds. Int J Stroke 2015;10:1297–1300.

46. Carter CJ. Genetic, Transcriptome, Proteomic, and Epidemiological Evidence for Blood-Brain Barrier Disruption and Polymicrobial Brain Invasion as Determinant Factors in Alzheimer’s Disease. J Alzheimers Dis Rep 2017;1:125–157.

47. Liang X, Liu C, Liu K, et al. Association and interaction of TOMM40 and PVRL2 with plasma amyloid-beta and Alzheimer’s disease among Chinese older adults: a population-based study. Neurobiol Aging 2022;113:143–151.

48. Chen YC, Chang SC, Lee YS, et al. TOMM40 Genetic Variants Cause Neuroinflammation in Alzheimer’s Disease. Int J Mol Sci 2023;24.

49. Ochoa D, Hercules A, Carmona M, et al. The next-generation Open Targets Platform: reimagined, redesigned, rebuilt. Nucleic Acids Res 2023;51:D1353–D1359.

50. Sollis E, Mosaku A, Abid A, et al. The NHGRI-EBI GWAS Catalog: knowledgebase and deposition resource. Nucleic Acids Res 2023;51:D977–D985.

51. Kuksa PP, Liu CL, Fu W, et al. Alzheimer’s Disease Variant Portal: A Catalog of Genetic Findings for Alzheimer’s Disease. J Alzheimers Dis 2022;86:461–477.

52. Agora [online]. Available at: https://agora.adknowledgeportal.org/. Accessed October 5, 2024.

53. Studies N-NWGoRiA, Chanock SJ, Manolio T, et al. Replicating genotype-phenotype associations. Nature 2007;447:655–660.

54. O’Connor LM, O’Connor BA, Lim SB, Zeng J, Lo CH. Integrative multi-omics and systems bioinformatics in translational neuroscience: A data mining perspective. J Pharm Anal 2023;13:836–850.

55. Landry LG, Ali N, Williams DR, Rehm HL, Bonham VL. Lack Of Diversity In Genomic Databases Is A Barrier To Translating Precision Medicine Research Into Practice. Health Aff (Millwood) 2018;37:780–785.

