## Supplementary material for "The Neurodegenerative Disease Knowledge Portal: Propelling Discovery Through the Sharing of Neurodegenerative Disease Genomic Resources": eTable 1

**eTable 1. Phenotypes encompassed by the Neurodegenerative Disease Knowledge Portal (NDKP)**

| Group | Phenotype |
| --- | --- |
| **Cerebrovascular MRI Traits** | Brain microbleeds (6 subtypes)<br>Cerebrovascular white matter hyperintensity volume<br>Fractional anisotropy in cerebral small vessel disease<br>Mean diffusivity in cerebral small vessel disease<br>White matter hyperintensities in cerebral small vessel disease |
| **Cognitive** | Cognitive performance<br>Intelligence<br>Memory performance<br>Subjective well-being<br>Verbal-numerical reasoning |
| **COVID-19** | COVID-19 (2 subtypes)<br>Hospitalized COVID-19 (2 subtypes)<br>Predicted COVID-19<br>Very severe respiratory confirmed COVID-19 (2 subtypes) |
| **Immunological** | Allergic disease (asthma, hay fever, or eczema)<br>Allergic rhinitis<br>Atopic dermatitis<br>Autoimmune Addison’s disease<br>Celiac disease<br>Graves' disease<br>Latent autoimmune diabetes in adults<br>Multiple sclerosis<br>Myasthenia gravis (3 subtypes)<br>Pollinosis<br>Psoriasis<br>Rheumatoid arthritis<br>Seropositive rheumatoid arthritis<br>Systemic lupus erythematosus (SLE) |
| **Metabolite** | Acetate<br>Acetoacetate<br>Alanine<br>Beta-hydroxybutyric acid<br>Calcium<br>Chloride<br>Citrate<br>Ferritin levels<br>Glutamine<br>Glycerol<br>Glycine<br>Glycoproteins<br>Histidine<br>Isoleucine<br>Lactate dehydrogenase (LDH) |

| Group | Phenotype |
| --- | --- |
| | Leucine<br>Phenylalanine<br>Phosphate<br>Phosphocholines<br>Phosphorus<br>Potassium<br>Pyruvate<br>Sodium<br>Total cholines<br>Tyrosine<br>Valine<br>Vitamin D levels |
| **Musculoskeletal** | Ankylosing spondylitis<br>Appendicular lean body mass<br>Back pain<br>Bone fracture<br>Bone mineral density (6 subtypes)<br>Femoral neck area<br>Grip strength (4 subtypes)<br>Heel quantitative ultrasound estimated bone mineral density (eBMD)<br>Hernia abdominopelvic cavity<br>Hip shape mode (10 subtypes)<br>Intertrochanteric-shaft area<br>Knee pain<br>Lumbar spine area<br>Osteoarthritis (12 subtypes)<br>Osteoporosis<br>Spinal canal stenosis<br>Total body lean mass<br>Total hip area<br>Total hip replacement<br>Total knee and-or hip replacement<br>Total knee replacement<br>Trochanter area<br>Usual walking pace |
| **Neurological** | Alzheimer's disease<br>Alzheimer’s disease family history (3 subtypes)<br>Ambidextrousness<br>Amyotrophic lateral sclerosis (ALS)<br>Carpal tunnel syndrome<br>Constipation in Parkinson's disease<br>Daytime sleepiness in Parkinson's disease<br>Dementia in Parkinson's disease<br>Depression in Parkinson's disease<br>Dyskinesias in Parkinson's disease<br>Epilepsy (generalized, with tonic-clonic seizures alone)<br>Focal epilepsy (2 subtypes)<br>Hoehn and Yahr score in Parkinson's disease<br>Hoehn and Yahr score of 3 or more in Parkinson's disease |

| Group | Phenotype |
|---|---|
| | Hyposmia in Parkinson's disease<br>Insomnia in Parkinson's disease<br>Late-onset Alzheimer's disease<br>Lewy body dementia<br>Mini-mental state examination in Parkinson's disease<br>Modified Schwab and England Activities of Daily Living Scale (SEADL) in Parkinson's disease (2 subtypes)<br>Montreal cognitive assessment in Parkinson's disease<br>Motor fluctuations in Parkinson's disease<br>Parkinson's disease<br>Parkinson's disease progression (3 subtypes)<br>Reaction time<br>REM sleep behavior disorder in Parkinson's disease<br>Restless legs syndrome in Parkinson's disease<br>Unified Parkinson's Disease Rating Scale OR the Movement Disorder Society revised (4 parts + total score) |
| **Psychiatric** | Acute reaction to stress<br>Alzheimer's disease family history<br>Attention-deficit hyperactivity disorder (ADHD)<br>Bipolar disorder<br>Depressed affect<br>Depression<br>Major depressive disorder (3 subtypes)<br>Neuroticism<br>Psychiatric disorders<br>Response to lithium treatment in bipolar disorder (2 subtypes)<br>Schizophrenia<br>Worry |
| **Sleep and Circadian** | Apnea-Hypopnea Index (AHI) (3 subtypes)<br>Average oxyhemoglobin desaturation per hypopnea event<br>Average oxyhemoglobin saturation during sleep<br>Chronotype (4 subtypes)<br>Daytime napping<br>Diurnal inactivity duration, rank-normalized<br>Ease of waking up<br>Excessive daytime sleepiness<br>Frequent insomnia symptoms<br>Least-active 5 hour timing, rank-normalized<br>Long sleep duration<br>Minimum oxyhemoglobin saturation (SpO2) during sleep<br>Most-active 10 hour timing, rank-normalized<br>Naps<br>Neck circumference (2 subtypes)<br>Number of sleep episodes, rank-normalized<br>Percentage of sleep with oxyhemoglobin saturation (SpO2) under 90%<br>Relative amplitude (2 subtypes)<br>Short sleep duration<br>Sleep duration (7 subtypes)<br>Sleep efficiency, rank-normalized |

| Group | Phenotype |
|---|---|
| | Sleep midpoint timing, rank-normalized<br>Snoring (2 subtypes) |
| **Stroke** | All intracranial hemorrhage<br>All ischemic stroke<br>Aneurysmal subarachnoid hemorrhage<br>Any stroke<br>Cardioembolic stroke<br>CCS causative cardio-aortic embolism major<br>CCS causative cryptogenic with cardioembolism minor<br>CCS causative incomplete and unclassified<br>CCS causative large artery atherosclerosis<br>CCS causative small artery occlusion<br>CCS causative undetermined<br>CCS phenotypic cardio-aortic embolism major<br>CCS phenotypic cryptogenic<br>CCS phenotypic cryptogenic large artery atherosclerosis major<br>CCS phenotypic cryptogenic small artery occlusion major<br>Intracerebral hemorrhage or small vessel ischemic stroke<br>Intracranial aneurysm<br>Lacunar stroke<br>Large artery stroke<br>Lobar intracerebral hemorrhage or small vessel ischemic stroke<br>Lobar intracranial hemorrhage<br>Modified Rankin scale score (6 subtypes)<br>Non-lobar intracerebral hemorrhage or small vessel ischemic stroke<br>Non-lobar intracranial hemorrhage<br>Post-stroke motor recovery<br>Small vessel stroke<br>TOAST cardio-aortic embolism<br>TOAST large artery atherosclerosis<br>TOAST other determined<br>TOAST other undetermined<br>TOAST small artery occlusion<br>Unruptured intracranial aneurysm |

Further details regarding the phenotypes captured by the Neurodegenerative Disease Knowledge Portal can be accessed online at: https://ndkp.hugeamp.org/research.html?pageid=kp_phenotypes&portal=ndkp. Abbreviations: CCS, Causative Classification System for Ischemic Stroke; REM, rapid eye movement; TOAST, Trial of Org 10172 in Acute Stroke Treatment.
